# Assessment of COVID-19 hospitalization risk during SARS-CoV-2 Omicron relative to Delta variant predominance, New York City, August 2021–January 2022

**DOI:** 10.1101/2022.07.15.22276814

**Authors:** Sharon K. Greene, Alison Levin-Rector, Elizabeth Luoma, Helly Amin, Emily McGibbon, Robert W. Mathes, Shama D. Ahuja

## Abstract

**Importance:** Assessing relative disease severity of SARS-CoV-2 variants in populations with varied vaccination and infection histories can help characterize emerging variants and support healthcare system preparedness.

**Objective:** To assess COVID-19 hospitalization risk for patients infected with Omicron (BA.1 and sublineages) compared with Delta SARS-CoV-2 variants.

**Design:** Observational cohort study.

**Setting:** New York City Department of Health and Mental Hygiene population-based COVID-19 disease registry, linked with laboratory results, immunization registry, and supplemental hospitalization data sources.

**Participants:** New York City residents with positive laboratory-based SARS-CoV-2 tests during August 2021–January 2022. A secondary analysis restricted to patients with whole-genome sequencing results, comprising 1%–18% of weekly confirmed cases.

**Exposures:** Diagnosis during periods when ≥98% of sequencing results were Delta (August–November 2021) or Omicron (January 2022). A secondary analysis defined variant exposure using patient-level sequencing results.

**Main outcomes and measures:** COVID-19 hospitalization, defined as a positive SARS-CoV-2 test 14 days before or 3 days after hospital admission.

**Results:** Among 646,852 persons with a positive laboratory-based SARS-CoV-2 test, hospitalization risk was lower for patients diagnosed when Omicron predominated (16,025/488,053, 3.3%) than when Delta predominated (8,268/158,799, 5.2%). In multivariable analysis adjusting for demographic characteristics and prior diagnosis and vaccination status, patients diagnosed when Omicron relative to Delta predominated had 0.72 (95% confidence interval [CI]: 0.63, 0.82) times the hospitalization risk. In a secondary analysis of 55,138 patients with sequencing results, hospitalization risk was similar for patients infected with Omicron (2,042/29,866, 6.8%) relative to Delta (1,780/25,272, 7.0%) and higher among those who received two mRNA vaccine doses (adjusted relative risk 1.64, 95% CI: 1.44, 1.87).

**Conclusions and relevance:** Illness severity was lower for patients diagnosed when Omicron (BA.1 and sublineages) relative to Delta predominated. This finding was consistent after adjusting for prior diagnosis and vaccination status, suggesting intrinsic virologic properties, not population-based immunity, accounted for the lower severity. A secondary analysis demonstrated collider bias from the sequencing sampling frame changing over time in ways associated with disease severity. Investing in representative data collection is necessary to avoid bias in assessing relative disease severity as new variants emerge, immunity wanes, and additional COVID-19 vaccines are administered.

## Introduction

Omicron (B.1.1.529 and BA lineages), a SARS-CoV-2 variant of concern, has caused less severe disease than prior variants in South Africa,^1,2^ Europe,^3-7^ and Canada.^8^ Similar findings have been reported in the U.S., based on national surveillance data and a large healthcare database,^9^ healthcare systems,^10-12^ and hospitals.^13,14^ Omicron severity assessments using population-based surveillance data linked to immunization registry data have been rare in the U.S.^15,16^ Few studies used patient-level whole-genome sequencing (WGS) results.^11,12,16,17^

Diminished disease severity could result from Omicron’s intrinsic virologic properties (e.g., lower replication competence in human lungs^18^), its emergence into populations with vaccination- and infection-induced immunity, or a combination thereof. Assessing variant severity in populations with varied vaccination and infection histories can help to clarify reasons for lower disease severity and support healthcare system preparedness.

In New York City (NYC), Delta (B.1.617.2 and AY lineages) became predominant (i.e., >50% of sequenced specimens) the week ending July 3, 2021, and Omicron became predominant the week ending December 18, 2021.^19^ Delta constituted 96% of sequencing results the first week of December 2021 and was swiftly displaced by Omicron, with 92% of sequencing results by the last week of December 2021.^19^ By the time Omicron was introduced into NYC, large percentages of the population were previously infected (a quarter of NYC residents were estimated to have been infected by mid-2020 before vaccine availability),^20,21^ and vaccinated (72% were fully vaccinated and 21% had received an additional dose as of December 18, 2021).^22^ During December 12, 2021–January 29, 2022, 1,047,428 confirmed and probable cases of COVID-19 were diagnosed among NYC residents.^23^ Given this surge in infections, we aimed to leverage population-based surveillance, WGS, and immunization registry data to assess relative disease severity for Omicron versus Delta infections, accounting for prior vaccination and diagnosis history.

## Methods

### Infection with Delta and Omicron variants

Clinical and commercial laboratories are required to report SARS-CoV-2 test results for New York State residents through the New York State Electronic Clinical Laboratory Reporting System (ECLRS).^24^ For some SARS-CoV-2 infections diagnosed among NYC residents, WGS is performed by the NYC Department of Health and Mental Hygiene (DOHMH) Public Health Laboratory, the Pandemic Response Laboratory,^25^ and other laboratories reporting through ECLRS.^26^ The Pandemic Response Laboratory reported most sequencing results during the study period, including 85% of sequencing results for patients diagnosed during January 2022. Variant assignment for sequence data from the Pandemic Response Laboratory and DOHMH were analyzed by the Public Health Laboratory with Pangolin version 4.0.6 and otherwise with Pangolin versions 3.1.8–3.1.20.

Using an observational cohort design, we defined a cohort of patients presumed infected with Delta as testing positive by laboratory-based molecular or antigen testing and diagnosed during August– November 2021 (≥98% of sequencing results were Delta during each week ending August 7–November 27, 2021).^19^ We defined a cohort of patients presumed infected with Omicron as diagnosed during January 2022 (≥99% of sequencing results were Omicron during each week ending January 8–29, 2022).^19^ Diagnosis date was defined as the specimen collection date of the first positive test within a 90-day period.

In a secondary analysis, we restricted to cases with sequencing results to eliminate uncertainty in the variant causing infection, at the expense of reduced sample size and representativeness (Supplement: eMethods). The weekly percentage of confirmed cases with sequencing results ranged from 1% of patients diagnosed during week ending January 1, 2022 at the peak of the Omicron wave to 18% during week ending December 4, 2021.^27^

### Hospitalization and death

COVID-19 hospitalizations and deaths are ascertained by importing and matching data from supplemental systems.^28^ Supplemental hospitalization data were obtained from emergency department syndromic surveillance, regional health information organizations, public hospitals, DOHMH’s electronic death registry system, and hospitals’ electronic health record systems.^28^ These systems do not capture all hospitalizations, and while all systems capture fact of hospitalization, the underlying cause is not necessarily available. Thus, people hospitalized because of COVID-19 illness could not be distinguished from people hospitalized because SARS-CoV-2 infection exacerbated an underlying condition or because SARS-CoV-2 infection was an incidental diagnosis.^29^ Incidental diagnoses were likely disproportionately common while Omicron predominated, given high infection prevalence.^30^

COVID-19 hospitalizations were defined as NYC residents whose positive SARS-CoV-2 test was within 14 days before or 3 days after hospital admission. COVID-19 deaths were defined as NYC residents with a positive SARS-CoV-2 test and (a) the cause-of-death on the death certificate was COVID-19 or similar, or b) COVID-19 was not a cause-of-death on the death certificate but the patient died within 30 days of COVID-19 diagnosis, and the death was not due to external causes such as injury.^28^ Sensitivity analyses applied more specific definitions (eMethods).

### Vaccination status

By matching with the NYC Citywide Immunization Registry, patients were assigned a vaccination status indicating the number of valid recorded doses (0–3) of an mRNA vaccine (BNT162b2 from Pfizer-BioNTech or mRNA-1273 from Moderna) received ≥14 days before diagnosis.^31^ We restricted third doses to those administered starting August 13, 2021, when the Advisory Committee on Immunization Practices recommended an additional dose after an initial series for eligible immunocompromised persons.^32^ Doses administered outside of New York State and by federal entities could have been missed, reducing the number of doses ascertained per patient.

### Diagnosis history

Prior diagnosis was defined as a positive laboratory-based molecular or antigen test >90 days before diagnosis with Delta or Omicron infection. Repeat positive tests are considered possible reinfections after 90 days, per the surveillance case definition.^33^ Prior diagnoses would have been missed for patients who resided outside of NYC when tested previously or who did not access testing, for instance during Spring 2020 when testing availability was limited.^28^

### Statistical analysis

The exposure of interest was infection with the Omicron or Delta variant, and the outcomes were hospitalization or death. Crude and adjusted relative risks (RRs) and 95% confidence intervals (CIs) were calculated using Poisson regression with robust error variance.^34^ Models adjusted for gender, age group (for hospitalizations in 10-year groupings as in Bager et al.:^3^ <10, 10–19, …, 80–89, ≥90 years; for deaths: same, except aggregating <30 year-olds), congregate setting residence (nursing home, jail, or prison; yes/no), and for community-dwelling residents, neighborhood poverty level (percent of residents based on census tract as of diagnosis with incomes below the federal poverty level, per the American Community Survey, 2015–2019).

To account for infection-induced immunity, we adjusted for prior COVID-19 diagnosis (yes/no) and, if yes, number of days since most recent prior diagnosis (91–179, 180–269, 270–359, 360–449, ≥450). Similarly, to account for vaccine-induced immunity, we adjusted for number of mRNA vaccine doses received ≥14 days before diagnosis (0–1, 2, or 3 doses) and number of days since 14 days after the most recent dose (<90, 90–179, 180–269, ≥270). Patients with 0 and 1 doses were aggregated for comparability with similar studies^3,12^ and because few patients received only 1 dose of the 2-dose primary series.

We included an interaction term for variant (Omicron vs. Delta) and vaccination status (0–1, 2, or 3 doses) and assessed risk of poor outcomes for patients diagnosed with Omicron relative to Delta infection within each vaccination status stratum. As in Bager et al.,^3^ we assessed risk of poor outcomes for both Omicron and Delta infections by vaccination status relative to patients with Delta infection and 0–1 mRNA vaccine doses received ≥14 days before diagnosis. Finally, restricting to patients diagnosed with Omicron infection, we assessed risk of poor outcomes by vaccination status relative to patients with 0–1 mRNA vaccine doses received ≥14 days before diagnosis.

Data were extracted from the DOHMH COVID-19 surveillance database (Maven Disease Surveillance and Outbreak Management System; Conduent), on March 10, 2022. Analyses were conducted using SAS Enterprise Guide, version 7.1 (SAS Institute). This activity was deemed public health surveillance that is non-research by the DOHMH Institutional Review Board.

## Results

After applying study exclusion criteria, the eligible population included 158,799 NYC residents diagnosed during August–November 2021 when Delta predominated and 488,053 persons diagnosed during January 2022 when Omicron predominated (Figure 1, Table 1). Of 21,023 persons diagnosed during August–November 2021 and with a Delta sequencing result, lineages constituting ≥5% of sequencing results were AY.103 (17%), B.1.617.2 (14%), AY.3 (10%), AY.44 (9%), AY.25 (7%), and AY.25.1 (6%). Of 19,274 persons diagnosed during January 2022 and with an Omicron sequencing result, lineages constituting ≥5% of sequencing results were BA.1.1 (42%), BA.1 (39%), BA.1.15 (6%), and BA.1.17.2 (6%); BA.1 and sublineages constituted 95.5% of sequencing results, with BA.2 and sublineages constituting the remaining 0.5%.

**Figure 1.**
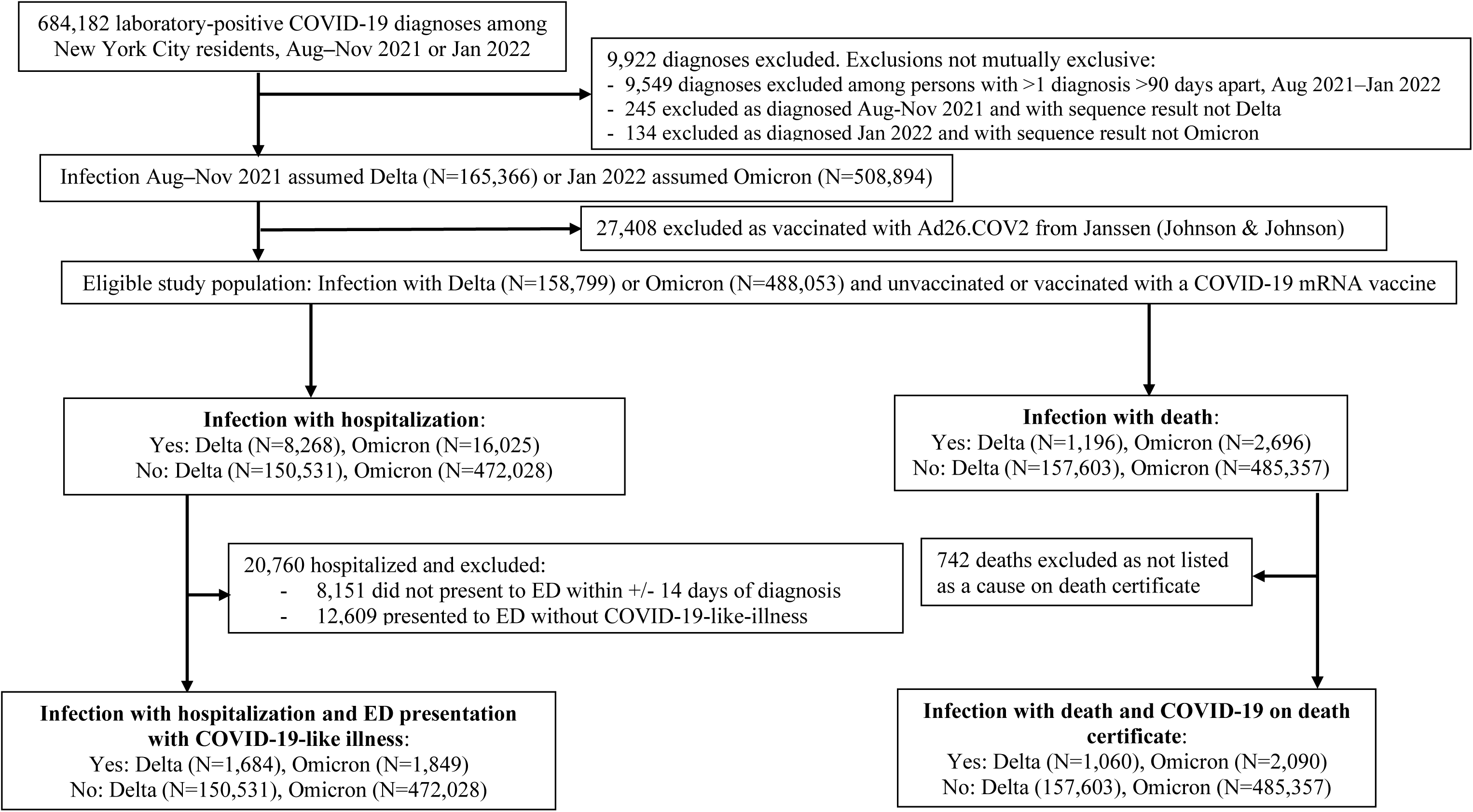
Eligibility for analysis of COVID-19 cases diagnosed during periods of Omicron or Delta predominance.

**Table 1.**
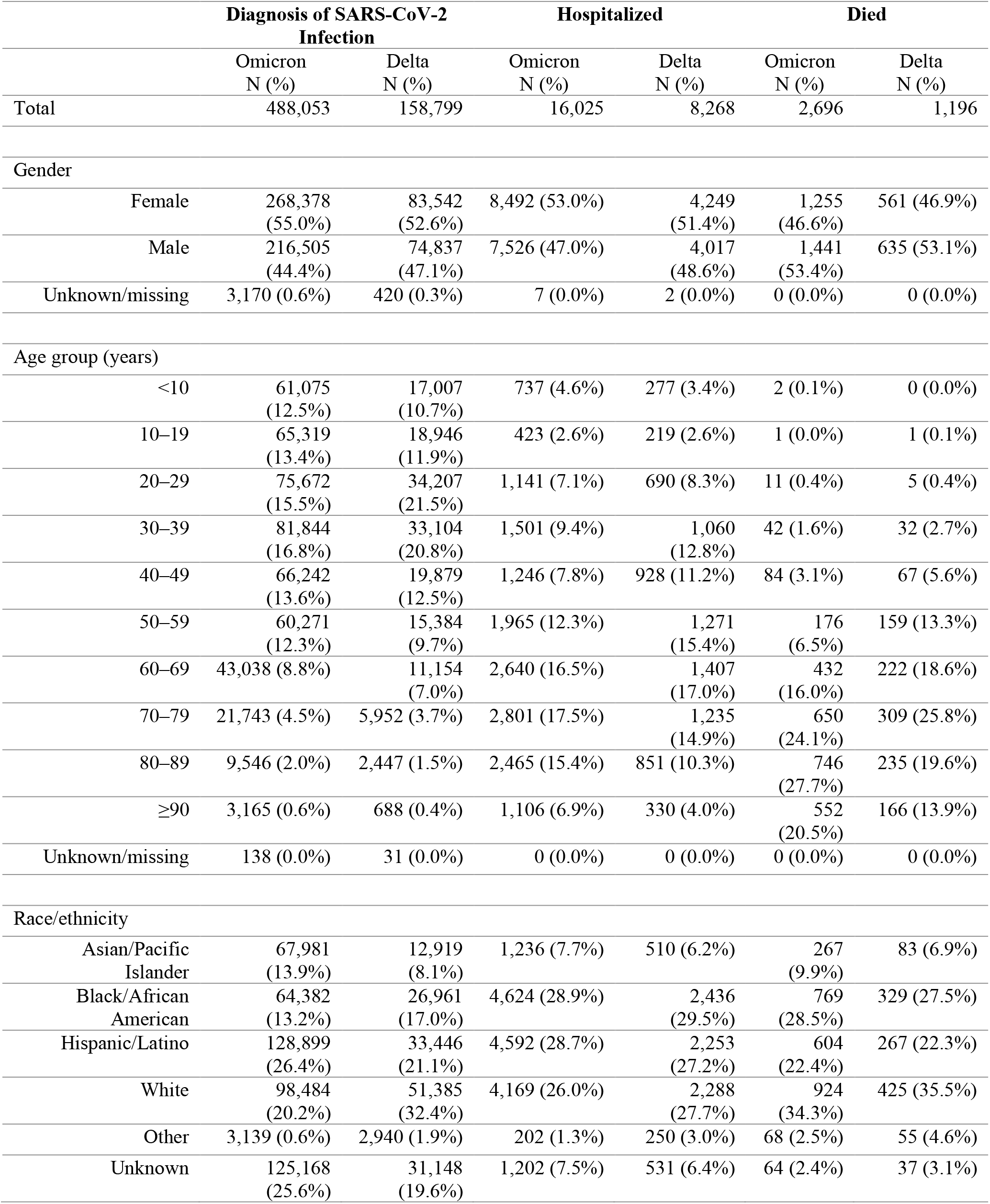
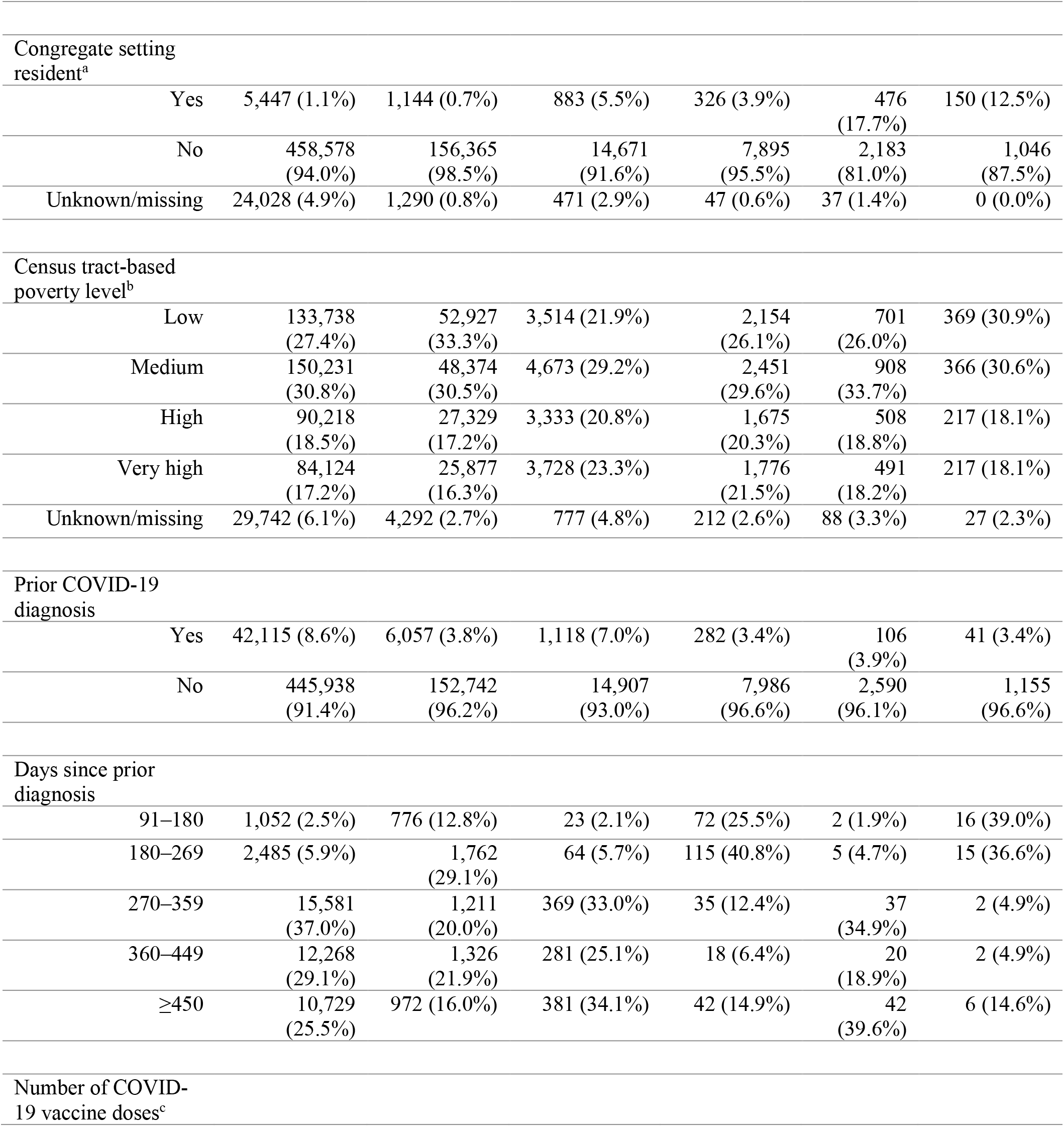
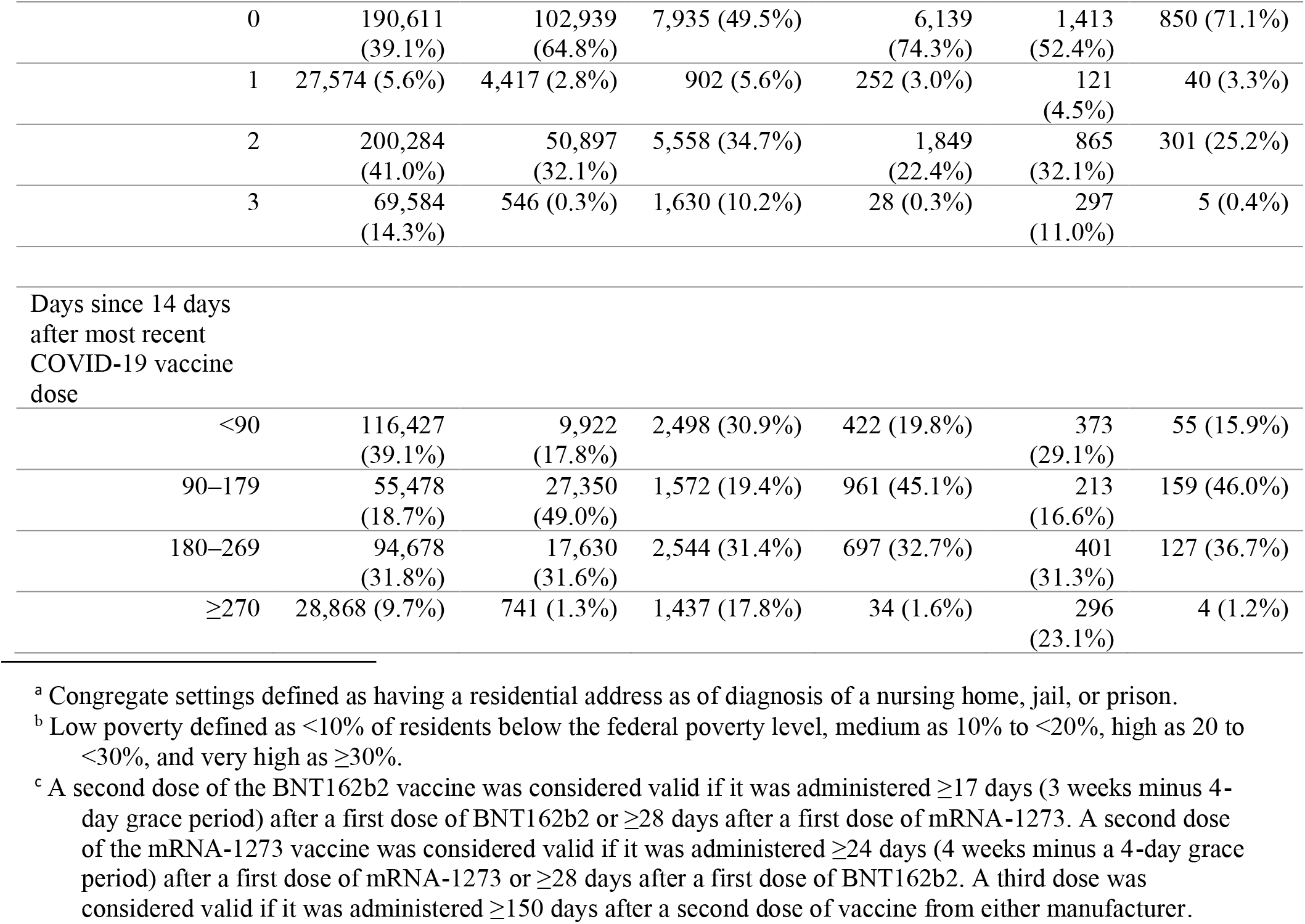
Characteristics of New York City residents diagnosed with COVID-19 during periods of Omicron (January 2022) and Delta (August–November 2021) variant predominance.

|  | Diagnosis of SARS-CoV-2 Infection |  | Hospitalized |  | Died |  |
| --- | --- | --- | --- | --- | --- | --- |
|  | Omicron<br>N (%) | Delta<br>N (%) | Omicron<br>N (%) | Delta<br>N (%) | Omicron<br>N (%) | Delta<br>N (%) |
| Total | 488,053 | 158,799 | 16,025 | 8,268 | 2,696 | 1,196 |
| Gender |  |  |  |  |  |  |
| Female | 268,378<br>(55.0%) | 83,542<br>(52.6%) | 8,492 (53.0%) | 4,249<br>(51.4%) | 1,255<br>(46.6%) | 561 (46.9%) |
| Male | 216,505<br>(44.4%) | 74,837<br>(47.1%) | 7,526 (47.0%) | 4,017<br>(48.6%) | 1,441<br>(53.4%) | 635 (53.1%) |
| Unknown/missing | 3,170 (0.6%) | 420 (0.3%) | 7 (0.0%) | 2 (0.0%) | 0 (0.0%) | 0 (0.0%) |
| Age group (years) |  |  |  |  |  |  |
| <10 | 61,075<br>(12.5%) | 17,007<br>(10.7%) | 737 (4.6%) | 277 (3.4%) | 2 (0.1%) | 0 (0.0%) |
| 10–19 | 65,319<br>(13.4%) | 18,946<br>(11.9%) | 423 (2.6%) | 219 (2.6%) | 1 (0.0%) | 1 (0.1%) |
| 20–29 | 75,672<br>(15.5%) | 34,207<br>(21.5%) | 1,141 (7.1%) | 690 (8.3%) | 11 (0.4%) | 5 (0.4%) |
| 30–39 | 81,844<br>(16.8%) | 33,104<br>(20.8%) | 1,501 (9.4%) | 1,060<br>(12.8%) | 42 (1.6%) | 32 (2.7%) |
| 40–49 | 66,242<br>(13.6%) | 19,879<br>(12.5%) | 1,246 (7.8%) | 928 (11.2%) | 84 (3.1%) | 67 (5.6%) |
| 50–59 | 60,271<br>(12.3%) | 15,384<br>(9.7%) | 1,965 (12.3%) | 1,271<br>(15.4%) | 176<br>(6.5%) | 159 (13.3%) |
| 60–69 | 43,038 (8.8%) | 11,154<br>(7.0%) | 2,640 (16.5%) | 1,407<br>(17.0%) | 432<br>(16.0%) | 222 (18.6%) |
| 70–79 | 21,743 (4.5%) | 5,952 (3.7%) | 2,801 (17.5%) | 1,235<br>(14.9%) | 650<br>(24.1%) | 309 (25.8%) |
| 80–89 | 9,546 (2.0%) | 2,447 (1.5%) | 2,465 (15.4%) | 851 (10.3%) | 746<br>(27.7%) | 235 (19.6%) |
| ≥90 | 3,165 (0.6%) | 688 (0.4%) | 1,106 (6.9%) | 330 (4.0%) | 552<br>(20.5%) | 166 (13.9%) |
| Unknown/missing | 138 (0.0%) | 31 (0.0%) | 0 (0.0%) | 0 (0.0%) | 0 (0.0%) | 0 (0.0%) |
| Race/ethnicity |  |  |  |  |  |  |
| Asian/Pacific Islander | 67,981<br>(13.9%) | 12,919<br>(8.1%) | 1,236 (7.7%) | 510 (6.2%) | 267<br>(9.9%) | 83 (6.9%) |
| Black/African American | 64,382<br>(13.2%) | 26,961<br>(17.0%) | 4,624 (28.9%) | 2,436<br>(29.5%) | 769<br>(28.5%) | 329 (27.5%) |
| Hispanic/Latino | 128,899<br>(26.4%) | 33,446<br>(21.1%) | 4,592 (28.7%) | 2,253<br>(27.2%) | 604<br>(22.4%) | 267 (22.3%) |
| White | 98,484<br>(20.2%) | 51,385<br>(32.4%) | 4,169 (26.0%) | 2,288<br>(27.7%) | 924<br>(34.3%) | 425 (35.5%) |
| Other | 3,139 (0.6%) | 2,940 (1.9%) | 202 (1.3%) | 250 (3.0%) | 68 (2.5%) | 55 (4.6%) |
| Unknown | 125,168<br>(25.6%) | 31,148<br>(19.6%) | 1,202 (7.5%) | 531 (6.4%) | 64 (2.4%) | 37 (3.1%) |
| Congregate setting resident<sup>a</sup> |  |  |  |  |  |  |
| Yes | 5,447 (1.1%) | 1,144 (0.7%) | 883 (5.5%) | 326 (3.9%) | 476 (17.7%) | 150 (12.5%) |
| No | 458,578 (94.0%) | 156,365 (98.5%) | 14,671 (91.6%) | 7,895 (95.5%) | 2,183 (81.0%) | 1,046 (87.5%) |
| Unknown/missing | 24,028 (4.9%) | 1,290 (0.8%) | 471 (2.9%) | 47 (0.6%) | 37 (1.4%) | 0 (0.0%) |
| Census tract-based poverty level<sup>b</sup> |  |  |  |  |  |  |
| Low | 133,738 (27.4%) | 52,927 (33.3%) | 3,514 (21.9%) | 2,154 (26.1%) | 701 (26.0%) | 369 (30.9%) |
| Medium | 150,231 (30.8%) | 48,374 (30.5%) | 4,673 (29.2%) | 2,451 (29.6%) | 908 (33.7%) | 366 (30.6%) |
| High | 90,218 (18.5%) | 27,329 (17.2%) | 3,333 (20.8%) | 1,675 (20.3%) | 508 (18.8%) | 217 (18.1%) |
| Very high | 84,124 (17.2%) | 25,877 (16.3%) | 3,728 (23.3%) | 1,776 (21.5%) | 491 (18.2%) | 217 (18.1%) |
| Unknown/missing | 29,742 (6.1%) | 4,292 (2.7%) | 777 (4.8%) | 212 (2.6%) | 88 (3.3%) | 27 (2.3%) |
| Prior COVID-19 diagnosis |  |  |  |  |  |  |
| Yes | 42,115 (8.6%) | 6,057 (3.8%) | 1,118 (7.0%) | 282 (3.4%) | 106 (3.9%) | 41 (3.4%) |
| No | 445,938 (91.4%) | 152,742 (96.2%) | 14,907 (93.0%) | 7,986 (96.6%) | 2,590 (96.1%) | 1,155 (96.6%) |
| Days since prior diagnosis |  |  |  |  |  |  |
| 91–180 | 1,052 (2.5%) | 776 (12.8%) | 23 (2.1%) | 72 (25.5%) | 2 (1.9%) | 16 (39.0%) |
| 180–269 | 2,485 (5.9%) | 1,762 (29.1%) | 64 (5.7%) | 115 (40.8%) | 5 (4.7%) | 15 (36.6%) |
| 270–359 | 15,581 (37.0%) | 1,211 (20.0%) | 369 (33.0%) | 35 (12.4%) | 37 (34.9%) | 2 (4.9%) |
| 360–449 | 12,268 (29.1%) | 1,326 (21.9%) | 281 (25.1%) | 18 (6.4%) | 20 (18.9%) | 2 (4.9%) |
| ≥450 | 10,729 (25.5%) | 972 (16.0%) | 381 (34.1%) | 42 (14.9%) | 42 (39.6%) | 6 (14.6%) |
| Number of COVID-19 vaccine doses<sup>c</sup> |  |  |  |  |  |  |

<sup>a</sup> Congregate settings defined as having a residential address as of diagnosis of a nursing home, jail, or prison.
<sup>b</sup> Low poverty defined as <10% of residents below the federal poverty level, medium as 10% to <20%, high as 20 to <30%, and very high as ≥30%.
<sup>c</sup> A second dose of the BNT162b2 vaccine was considered valid if it was administered ≥17 days (3 weeks minus 4-day grace period) after a first dose of BNT162b2 or ≥28 days after a first dose of mRNA-1273. A second dose of the mRNA-1273 vaccine was considered valid if it was administered ≥24 days (4 weeks minus a 4-day grace period) after a first dose of mRNA-1273 or ≥28 days after a first dose of BNT162b2. A third dose was considered valid if it was administered ≥150 days after a second dose of vaccine from either manufacturer.

|  |  |  |  |  |  |  |
| --- | --- | --- | --- | --- | --- | --- |
| 0 | 190,611<br>(39.1%) | 102,939<br>(64.8%) | 7,935 (49.5%) | 6,139<br>(74.3%) | 1,413<br>(52.4%) | 850 (71.1%) |
| 1 | 27,574 (5.6%) | 4,417 (2.8%) | 902 (5.6%) | 252 (3.0%) | 121<br>(4.5%) | 40 (3.3%) |
| 2 | 200,284<br>(41.0%) | 50,897<br>(32.1%) | 5,558 (34.7%) | 1,849<br>(22.4%) | 865<br>(32.1%) | 301 (25.2%) |
| 3 | 69,584<br>(14.3%) | 546 (0.3%) | 1,630 (10.2%) | 28 (0.3%) | 297<br>(11.0%) | 5 (0.4%) |
| Days since 14 days<br>after most recent<br>COVID-19 vaccine<br>dose |  |  |  |  |  |  |
| <90 | 116,427<br>(39.1%) | 9,922<br>(17.8%) | 2,498 (30.9%) | 422 (19.8%) | 373<br>(29.1%) | 55 (15.9%) |
| 90–179 | 55,478<br>(18.7%) | 27,350<br>(49.0%) | 1,572 (19.4%) | 961 (45.1%) | 213<br>(16.6%) | 159 (46.0%) |
| 180–269 | 94,678<br>(31.8%) | 17,630<br>(31.6%) | 2,544 (31.4%) | 697 (32.7%) | 401<br>(31.3%) | 127 (36.7%) |
| ≥270 | 28,868 (9.7%) | 741 (1.3%) | 1,437 (17.8%) | 34 (1.6%) | 296<br>(23.1%) | 4 (1.2%) |

Demographic characteristics were similar between cohorts; patients were predominantly female, 20–39 years-old, and residents of medium poverty areas (Table 1, eTable 1). Patients diagnosed during Omicron predominance were more likely to have been vaccinated with 1, 2, and 3 doses than patients infected during Delta predominance (Table 1), as expected because the Omicron wave occurred after the Delta wave, allowing more time for vaccine administration. As there was little temporal overlap between the Delta wave and third vaccine dose administrations,^22^ only 0.3% of patients diagnosed with Delta infection had received 3 doses (Table 1).

History of prior COVID-19 diagnosis was more common for patients diagnosed during Omicron (8.6%) than Delta predominance (3.8%) (Table 1). Study eligibility required patients to have had either Delta or Omicron infection but not both. Thus, with Omicron emerging after Delta, patients infected with Omicron had on average more time for infection-induced immunity to wane. The median number of days since prior diagnosis for Omicron reinfections was 367 (interquartile range: 317–467) and for Delta reinfections was 300 (interquartile range: 224–365).

Missingness for covariates included in regression analyses was negligible (≤1.5%), except variables depending on geocoding, i.e., congregate setting residence and neighborhood poverty level, had up to 6.1% missingness (Table 1). We conducted regression modeling using complete case analysis, assuming data were missing completely at random.

### Hospitalization and death

Of 488,053 NYC residents diagnosed during weeks when ≥99% of sequencing results were Omicron and presumed infected with Omicron, 16,025 (3.3%) were hospitalized, and 2,696 (0.6%) died. Of 158,799 persons diagnosed when ≥98% of sequencing results were Delta and presumed infected with Delta, 8,268 (5.2%) were hospitalized, and 1,196 (0.8%) died. Patients infected with Omicron relative to Delta had 0.72 (95% confidence interval [CI]: 0.63, 0.82) times the risk of hospitalization, adjusting for gender, age, congregate setting residence, neighborhood poverty level, prior COVID-19 diagnosis, time since prior COVID-19 diagnosis, number of mRNA vaccine doses received, and number of days since 14 days after the most recent vaccine dose (Figure 2, eTable 2). The point estimate for deaths among patients infected with Omicron relative to Delta was similar to that of hospitalizations but with wider uncertainty (adjusted relative risk [aRR] 0.81, 95% CI: 0.58, 1.13) (Figure 2, eTable 3). In sensitivity analyses using more specific outcome definitions with sparser observations, point estimates remained similar for severity of Omicron relative to Delta infection, with 0.70 (95% CI: 0.36, 1.34) times the adjusted risk of hospitalization with COVID-19-like illness presentation and 0.86 (95% CI: 0.54, 1.38) times the adjusted risk of death with COVID-19 indicated on the death certificate (eFigure 2).

**Figure 2:**
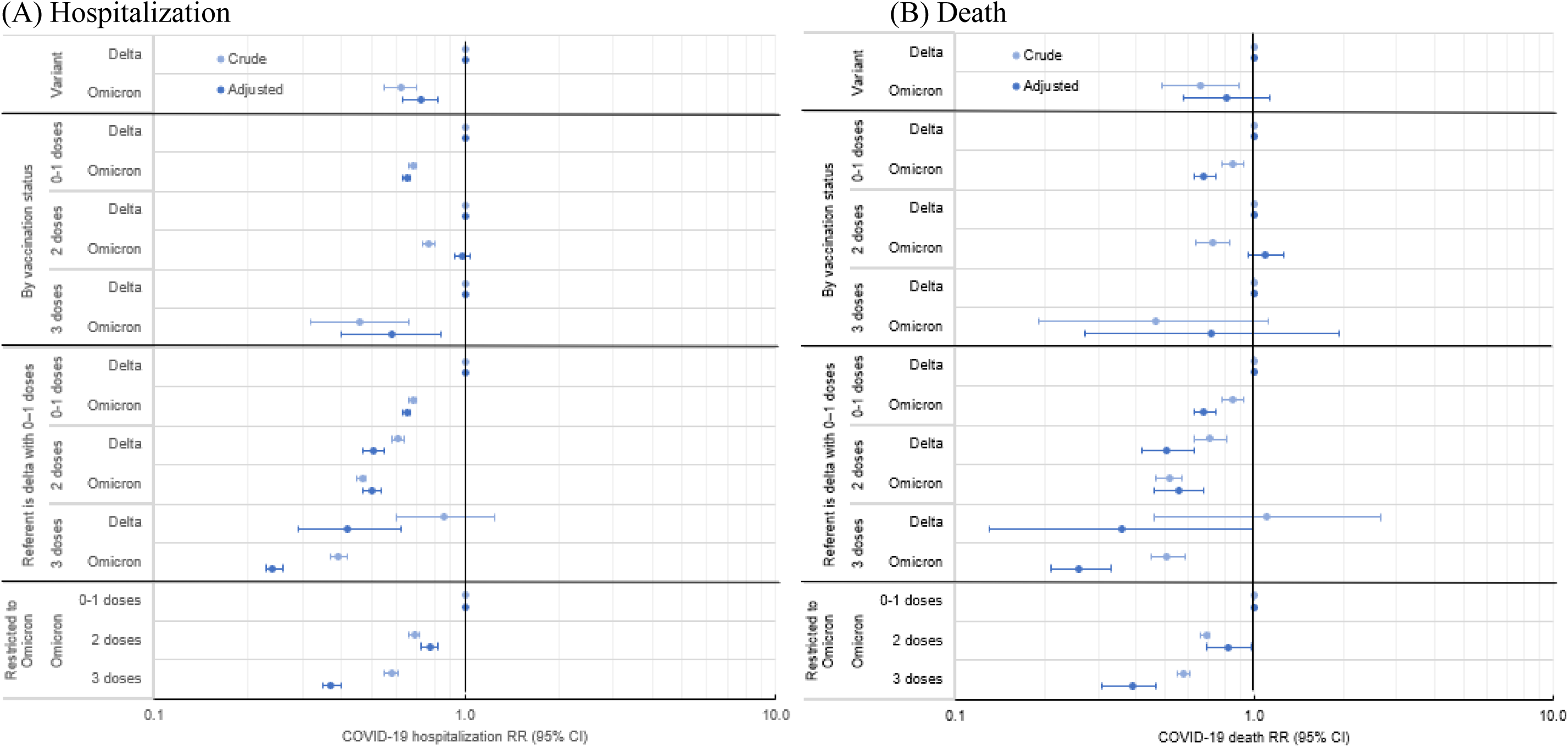
Relative risks of COVID-19 hospitalization and death by diagnosis during periods of Omicron compared with Delta predominance, overall and according to vaccination status, New York City, August 2021–January 2022.

Associations between poor outcomes and variant were modified by vaccination status; the interaction term for variant and vaccination status in adjusted models was p<0.0001 for all four outcomes (hospitalization, hospitalization with COVID-19-like illness presentation, death, and death with COVID-19 indicated on the death certificate). By vaccination status, the aRR of hospitalization for patients infected with Omicron relative to Delta was 0.65 (95% CI: 0.63, 0.67) among patients with 0–1 vaccine dose, 0.98 (95% CI: 0.93, 1.04) among those who received 2 doses, and 0.58 (95% CI: 0.40, 0.84) among those who received 3 doses (Figure 2, eTable 2). The pattern for all four outcomes was similar, in that severity for patients infected with Omicron relative to Delta was strongly reduced among patients who received 0–1 vaccine dose but less reduced or not reduced among those who received 2 doses (Figure 2, eFigure 2). Relative severity estimates among patients who received 3 doses had wide uncertainty given few patients infected with Delta had received 3 doses. However, restricting to patients infected with Omicron and relative to those with 0–1 doses, those who received 2 vaccine doses had 0.77 times the adjusted risk of hospitalization (95% CI: 0.72, 0.82) and 0.82 times the adjusted risk of death (95% CI: 0.69, 0.98); those who received 3 doses had 0.37 times the adjusted risk of hospitalization (95% CI: 0.35, 0.40) and 0.39 times the adjusted risk of death (95% CI: 0.31, 0.47) (Figure 2, eTables 2–3).

Furthermore, we assessed risk of poor outcomes by vaccination status relative to patients infected with Delta who received 0–1 vaccine dose. Risk of hospitalization after 2 doses was similar between patients with Omicron (aRR 0.50, 95% CI: 0.47, 0.54) and Delta infections (aRR 0.51, 95% CI: 0.47, 0.55), and risk of hospitalization after 3 vaccine doses was lower with Omicron (aRR 0.24, 95% CI: 0.23, 0.26) than with Delta (aRR 0.42, 95% CI: 0.29, 0.62) (eTable 2).

Results were not robust in secondary analyses restricting to patients with sequencing results. Of 29,866 NYC residents with Omicron sequencing results, 2,042 (6.8%) were hospitalized, and 446 (1.5%) died (eTable 1). Of 25,272 patients with Delta sequencing results, 1,780 (7.0%) were hospitalized, and 331 (1.3%) died. The aRR of hospitalization for patients infected with Omicron relative to Delta based on sequencing results was increased among those who received 2 doses (aRR 1.64, 95% CI: 1.44, 1.87) (eFigure 3). This finding of increased relative severity for Omicron infections was inconsistent with both the primary analysis and prior literature and likely reflects a bias in which specimens from severely ill patients were disproportionately selected for sequencing when Omicron predominated.

## Discussion

NYC residents diagnosed when Omicron (BA.1 and sublineages) relative to Delta predominated had lower hospitalization risk, as has consistently been reported. This finding was consistent in both crude and adjusted analyses controlling for prior diagnosis and vaccination status, and we found lower hospitalization and death risk among patients with 0–1 vaccine dose, suggesting Omicron’s diminished disease severity is likely more attributable to intrinsic virologic properties than to prior population-based immunity. Among patients with 2 vaccine doses, those infected with Omicron had similar hospitalization and death risk as those infected with Delta, possibly reflecting Omicron’s increased ability to evade vaccine-induced immunity.^35,36^ Analyses did not include persons only testing positive using at-home rapid antigen tests, which became more widely available starting mid-December 2021 while Omicron predominated.^37,38^ If at-home tests were used differentially by persons with milder illness, then patients diagnosed while Omicron predominated would have been biased toward severe illness. Thus, our estimates are likely conservative, and Omicron could be even less severe relative to Delta than we report. Despite reduced relative severity, the absolute numbers of COVID-19 hospitalizations and deaths were higher when Omicron predominated given the volume of Omicron infections.

We analyzed 646,852 persons with Delta or Omicron infection, >3 times as large a study population as the 188,980 persons included in a similar, Danish analysis.^3^ Findings were consistent, except the Danish analysis showed that among those who received 2 vaccine doses, patients infected with Omicron relative to Delta had lower risk of hospitalization (aRR 0.71, 95% CI: 0.60, 0.86), while our study showed no difference (aRR 0.98, 95% CI: 0.93, 1.04). Reasons for this discrepancy are unclear, although the confounder adjustment set was different (we adjusted for congregate setting residence, neighborhood poverty level, time since vaccination, and time since prior COVID-19 diagnosis, while the Danish analysis adjusted for presence of comorbidities, which was unavailable for our cohort), and vaccination information could have been more complete from the Danish national registry.

In addition to the large population, other study strengths included assessment of multiple poor outcomes, as well as a demonstration using real-world surveillance data of limitations of restricting to patients with WGS results to assess poor outcomes. A secondary analysis implausibly suggested higher disease severity for Omicron relative to Delta infections among those who received 2 vaccine doses. Only 1% of confirmed cases had sequencing results at the Omicron wave’s peak, when the Pandemic Response Laboratory and Public Health Laboratory prioritized specimens with low cycle threshold values for sequencing. Conditioning analyses on patients seeking voluntary testing and having specimens selected for sequencing, the probabilities of which changed over time concurrent with changing variant predominance, likely induced collider bias.^39,40^ This underscores the importance of a representative WGS sampling frame that does not change over time in ways associated with disease severity. Outcome comparisons for patients with sequencing results should be interpreted cautiously if not diagnosed closely in time. Sample size calculations for variant surveillance should consider not only how many sequences are needed to detect new variants or monitor sequence prevalence,^41^ but also the much larger number needed to assess relative risk of hospitalization and death across variants, stratifying by vaccination status, to support healthcare capacity planning.

SARS-CoV-2 variant surveillance exemplifies investments in public health data and informatics. Our large, population-based study relied on DOHMH capacity to link confidential, identifiable patient data in the COVID-19 disease registry with laboratory data for sequencing results, with the immunization registry for vaccination status, with past cases for diagnosis history, with supplemental data sources for hospitalizations, and with the vital statistics registry for deaths. Capacity to access and link data sources varies across U.S. health departments. Although we did not have access to all relevant variables (e.g., comorbidities, therapeutics receipt, intensive care unit admission, and mechanical ventilation), such data are more readily available within healthcare systems.^10^ Strengthening linkages between healthcare and public health data systems will enable continued assessment of relative disease severity as new variants and subvariants (e.g., BA.4, BA.5) emerge, immunity from prior infections and vaccination wanes, and additional COVID-19 vaccines are administered. Among NYC residents diagnosed with Omicron infection and relative to those with 0–1 mRNA vaccine dose, those who received 3 doses had strongly reduced risk of hospitalization and death, supporting continued efforts to ensure up-to-date vaccination coverage.

## Data Availability

Line-level data are not publicly available in accordance with patient confidentiality and privacy laws. Publicly available data are linked below.

https://www1.nyc.gov/site/doh/covid/covid-19-data.page

## Acknowledgments

The authors thank all DOHMH staff serving in the Surveillance and Epidemiology Section, Laboratory Emergency Response Group, and Vaccine Operations Center of the NYC Incident Command System. We thank Nang TT Kyaw for contributions to the literature review and analysis and the Citywide Immunization Registry Research and Evaluation Team as well as Jennifer Baumgartner, Katelynn Devinney, Kathleen H. Reilly, and Carolyn Chang for contributions to data management. We also thank Rebecca Kahn and Keya Joshi (Center for Communicable Disease Dynamics, Harvard TH Chan School of Public Health) for clarifying discussions about methods and limitations.

## Disclaimer

The findings and conclusions in this report are those of the authors and do not necessarily represent the official position of the New York City Department of Health and Mental Hygiene.

## Conflict of Interest Disclosures

None.

## Funding/Support

The authors received no specific funding for this work beyond their usual salaries. Dr. Greene was supported by the Public Health Emergency Preparedness Cooperative Agreement (grant No. NU90TP922035-03-03), funded by the US Centers for Disease Control and Prevention (CDC). Ms. Levin-Rector was supported by ELC CARES (grant No. NU50CK000517-01-09), funded by CDC.

## Role of the Funder/Sponsor

This manuscript completed clearance review and approval processes at NYC DOHMH prior to submission. This work received no specific funding; thus, there were no funding organizations with any role in the design and conduct of the study; collection, management, analysis, and interpretation of the data; preparation of the manuscript; or decision to submit the manuscript for publication.

## Notes

### Competing Interest Statement

The authors have declared no competing interest.

### Author Declarations

The Institutional Review Board of the New York City Department of Health and Mental Hygiene gave ethical approval for this work.

