## Supplement for "Assessment of COVID-19 hospitalization risk during SARS-CoV-2 Omicron relative to Delta variant predominance, New York City, August 2021–January 2022"

**eMethods**

**eTable 1.** Characteristics of New York City residents with Delta and Omicron SARS-CoV-2 infections based on sequencing result, diagnosed July 2021–January 2022.

**eTable 2**. Relative risks of hospitalization by diagnosis with COVID-19 during periods of Omicron compared with Delta predominance, overall and according to vaccination status, New York City, August 2021–January 2022.

**eTable 3.** Relative risks of death by diagnosis with COVID-19 during periods of Omicron compared with Delta predominance, overall and according to vaccination status, New York City, August 2021–January 2022.

**eFigure 1.** Eligibility for secondary analysis of Delta and Omicron infections based on whole-genome sequencing result.

**eFigure 2.** Relative risks of (A) hospitalization with COVID-19-like illness presentation and (B) death with COVID-19 indicated on the death certificate, by diagnosis during periods of Omicron compared with Delta predominance overall and according to vaccination status, New York City, August 2021–January 2022.

**eFigure 3.** Relative risks of COVID-19 hospitalization and death for patients with Omicron compared with Delta sequencing results, overall and according to vaccination status, New York City, July 2021–January 2022.

This supplemental material has been provided by the authors to give readers additional information about their work.

### eMethods.

#### Sensitivity analyses

For a more specific exposure definition, we alternatively identified patients diagnosed during July 1, 2021–January 31, 2022 and with sequencing results indicating infection with Delta (n=25,272) or Omicron (n=29,866) variant (eFigure 1).

For a more specific hospitalization definition, we restricted to the subset of COVID-19 hospitalizations for which, based on syndromic surveillance, patients had presented to the emergency department with COVID-19-like illness within +/- 14 days of diagnosis. COVID-19-like illness was defined as chief complaint or ICD-10 codes for pneumonia or influenza-like illness (fever, cough, sore throat, and/or respiratory illness), and without regard to the COVID-19 ICD-10 code.

For a more specific deaths definition, we restricted to the subset with COVID-19 indicated on the death certificate.

#### Exclusions

Exclusion criteria included patients with >1 diagnosis >90 days apart during the study period, such that each eligible patient had only Delta or Omicron infection (Figure 1). Additionally, on December 16, 2021, the Advisory Committee on Immunization Practices preferentially recommended the mRNA COVID-19 vaccines.[1] The small percentage of study participants whose first COVID-19 vaccine dose was Ad26.COV2 from Janssen (Johnson & Johnson) were excluded (4% [27,408/674,260], Figure 1). With the operational benefit of requiring only one dose, this vaccine had been differentially administered to certain populations, e.g., homebound seniors.[2] These vaccine recipients likely disproportionately had underlying conditions associated with COVID-19 hospitalization and death, and including them without accounting for these unobserved confounders could have biased results.

1. Oliver SE, Wallace M, See I, et al. Use of the Janssen (Johnson & Johnson) COVID-19 vaccine: updated interim recommendations from the Advisory Committee on Immunization Practices - United States, December 2021. MMWR Morb Mortal Wkly Rep. 2022;71(3):90-95.
2. The City of New York - Office of the Mayor. Vaccine for all: City begins vaccination for homebound New Yorkers. Mar 4, 2021. https://www1.nyc.gov/office-of-the-mayor/news/155-21/vaccine-all-city-begins-vaccination-homebound-new-yorkers. Accessed Feb 2, 2022.

**eTable 1. Characteristics of New York City residents with Delta and Omicron SARS-CoV-2 infections based on sequencing result, diagnosed July 2021–January 2022.**

| | Diagnosis of SARS-CoV-2 Infection | | Hospitalized | | Died | |
|---|---|---|---|---|---|---|
| | Omicron N (%) | Delta N (%) | Omicron N (%) | Delta N (%) | Omicron N (%) | Delta N (%) |
| Total | 29,866 | 25,272 | 2,042 | 1,780 | 446 | 331 |
| Gender | | | | | | |
| Female | 16,182 (54.2%) | 13,273 (52.5%) | 1,065 (52.2%) | 925 (52.0%) | 181 (40.6%) | 152 (45.9%) |
| Male | 13,250 (44.4%) | 11,957 (47.3%) | 977 (47.8%) | 854 (48.0%) | 265 (59.4%) | 179 (54.1%) |
| Unknown/missing | 434 (1.5%) | 42 (0.2%) | 0 (0.0%) | 1 (0.1%) | 0 (0.0%) | 0 (0.0%) |
| Age group (years) | | | | | | |
| <10 | 2,998 (10.0%) | 2,612 (10.3%) | 126 (6.2%) | 43 (2.4%) | 0 (0.0%) | 0 (0.0%) |
| 10–19 | 3,049 (10.2%) | 2,906 (11.5%) | 70 (3.4%) | 48 (2.7%) | 0 (0.0%) | 0 (0.0%) |
| 20–29 | 5,771 (19.3%) | 5,087 (20.1%) | 158 (7.7%) | 133 (7.5%) | 7 (1.6%) | 1 (0.3%) |
| 30–39 | 5,766 (19.3%) | 5,607 (22.2%) | 219 (10.7%) | 225 (12.6%) | 13 (2.9%) | 12 (3.6%) |
| 40–49 | 3,963 (13.3%) | 3,391 (13.4%) | 142 (7.0%) | 242 (13.6%) | 19 (4.3%) | 24 (7.3%) |
| 50–59 | 3,457 (11.6%) | 2,606 (10.3%) | 226 (11.1%) | 285 (16.0%) | 40 (9.0%) | 43 (13.0%) |
| 60–69 | 2,639 (8.8%) | 1,755 (6.9%) | 324 (15.9%) | 298 (16.7%) | 77 (17.3%) | 71 (21.5%) |
| 70–79 | 1,270 (4.3%) | 828 (3.3%) | 354 (17.3%) | 262 (14.7%) | 100 (22.4%) | 78 (23.6%) |
| 80–89 | 699 (2.3%) | 369 (1.5%) | 305 (14.9%) | 176 (9.9%) | 123 (27.6%) | 59 (17.8%) |
| ≥90 | 254 (0.9%) | 111 (0.4%) | 118 (5.8%) | 68 (3.8%) | 67 (15.0%) | 43 (13.0%) |
| Unknown/missing | 0 (0.0%) | 0 (0.0%) | 0 (0.0%) | 0 (0.0%) | 0 (0.0%) | 0 (0.0%) |
| Race/ethnicity | | | | | | |
| Asian/Pacific Islander | 3,648 (12.2%) | 2,048 (8.1%) | 168 (8.2%) | 118 (6.6%) | 43 (9.6%) | 20 (6.0%) |
| Black/African American | 4,816 (16.1%) | 5,014 (19.8%) | 690 (33.8%) | 592 (33.3%) | 138 (30.9%) | 111 (33.5%) |
| Hispanic/Latino | 7,653 (25.6%) | 5,967 (23.6%) | 490 (24.0%) | 466 (26.2%) | 96 (21.5%) | 77 (23.3%) |
| White | 6,712 (22.5%) | 7,280 (28.8%) | 430 (21.1%) | 432 (24.3%) | 142 (31.8%) | 102 (30.8%) |
| Other | 200 (0.7%) | 537 (2.1%) | 23 (1.1%) | 58 (3.3%) | 11 (2.5%) | 12 (3.6%) |

| | | | | | | |
|---|---|---|---|---|---|---|
| Unknown | 6,837 (22.9%) | 4,426 (17.5%) | 241 (11.8%) | 114 (6.4%) | 16 (3.6%) | 9 (2.7%) |
| Congregate setting resident[a] | | | | | | |
| Yes | 347 (1.2%) | 119 (0.5%) | 104 (5.1%) | 59 (3.3%) | 60 (13.5%) | 36 (10.9%) |
| No | 28,628 (95.9%) | 24,951 (98.7%) | 1,856 (90.9%) | 1,713 (96.2%) | 379 (85.0%) | 295 (89.1%) |
| Unknown/missing | 891 (3.0%) | 202 (0.8%) | 82 (4.0%) | 8 (0.4%) | 7 (1.6%) | 0 (0.0%) |
| Census tract-based poverty level[b] | | | | | | |
| Low | 8,801 (29.5%) | 7,816 (30.9%) | 343 (16.8%) | 371 (20.8%) | 89 (20.0%) | 77 (23.3%) |
| Medium | 9,254 (31.0%) | 7,830 (31.0%) | 708 (34.7%) | 562 (31.6%) | 169 (37.9%) | 103 (31.1%) |
| High | 5,358 (17.9%) | 4,538 (18.0%) | 386 (18.9%) | 380 (21.3%) | 77 (17.3%) | 56 (16.9%) |
| Very high | 5,094 (17.1%) | 4,255 (16.8%) | 498 (24.4%) | 420 (23.6%) | 101 (22.6%) | 87 (26.3%) |
| Unknown/missing | 1,359 (4.6%) | 833 (3.3%) | 107 (5.2%) | 47 (2.6%) | 10 (2.2%) | 8 (2.4%) |
| Prior COVID-19 diagnosis | | | | | | |
| Yes | 2,137 (7.2%) | 1,251 (5.0%) | 130 (6.4%) | 24 (1.3%) | 19 (4.3%) | 3 (0.9%) |
| No | 27,729 (92.8%) | 24,021 (95.0%) | 1,912 (93.6%) | 1,756 (98.7%) | 427 (95.7%) | 328 (99.1%) |
| Days since prior diagnosis | | | | | | |
| 91–180 | 153 (7.2%) | 99 (7.9%) | 12 (9.2%) | 10 (41.7%) | 3 (15.8%) | 2 (66.7%) |
| 180–269 | 166 (7.8%) | 218 (17.4%) | 10 (7.7%) | 8 (33.3%) | 1 (5.3%) | 0 (0.0%) |
| 270–359 | 764 (35.8%) | 203 (16.2%) | 47 (36.2%) | 2 (8.3%) | 3 (15.8%) | 0 (0.0%) |
| 360–449 | 533 (24.9%) | 637 (50.9%) | 30 (23.1%) | 1 (4.2%) | 2 (10.5%) | 0 (0.0%) |
| ≥450 | 521 (24.4%) | 93 (7.4%) | 31 (23.8%) | 3 (12.5%) | 10 (52.6%) | 1 (33.3%) |
| Number of COVID-19 vaccine doses | | | | | | |
| 0 | 10,863 (36.4%) | 16,006 (63.3%) | 1,105 (54.1%) | 1,405 (78.9%) | 271 (60.8%) | 258 (77.9%) |
| 1 | 1,502 (5.0%) | 680 (2.7%) | 87 (4.3%) | 53 (3.0%) | 21 (4.7%) | 10 (3.0%) |

[a] Congregate settings defined as having a residential address as of diagnosis of a nursing home, jail, or prison.
[b] Low poverty defined as <10% of residents below the federal poverty level, medium as 10% to <20%, high as 20 to <30%, and very high as ≥30%.

| | | | | | | |
|---|---|---|---|---|---|---|
| 2 | 13,129 (44.0%) | 8,443 (33.4%) | 693 (33.9%) | 310 (17.4%) | 119 (26.7%) | 58 (17.5%) |
| 3 | 4,372 (14.6%) | 143 (0.6%) | 157 (7.7%) | 12 (0.7%) | 35 (7.8%) | 5 (1.5%) |
| Days since 14 days after most recent COVID-19 vaccine dose | | | | | | |
| <90 | 6,906 (36.3%) | 1,788 (19.3%) | 263 (28.1%) | 82 (21.9%) | 46 (26.3%) | 17 (23.3%) |
| 90–179 | 3,295 (17.3%) | 3,889 (42.0%) | 180 (19.2%) | 133 (35.5%) | 24 (13.7%) | 27 (37.0%) |
| 180–269 | 6,735 (35.4%) | 3,245 (35.0%) | 350 (37.4%) | 141 (37.6%) | 63 (36.0%) | 27 (37.0%) |
| ≥270 | 1,967 (10.4%) | 314 (3.4%) | 141 (15.0%) | 16 (4.3%) | 42 (24.0%) | 1 (1.4%) |

**eTable 2. Relative risks of hospitalization by diagnosis with COVID-19 during periods of Omicron compared with Delta predominance, overall and according to vaccination status, New York City, August 2021–January 2022.**

| | Hospitalized | Not hospitalized | Crude RR (95% CI) | p value | Adjusted RR (95% CI) | p value |
|---|---|---|---|---|---|---|
| **Variant** | | | | | | |
| Delta | 8,268/158,799 (5.2%) | 150,531/158,799 (94.8%) | 1 (ref) | | 1 (ref) | |
| Omicron | 16,025/488,053 (3.3%) | 472,028/488,053 (96.7%) | 0.62 (0.55, 0.70) | <.001 | 0.72 (0.63, 0.82) | <.001 |
| **By vaccination status** | | | | | | |
| None or only one dose | | | | <.001 | | <.001 |
| Delta | 6,391/107,356 (6.0%) | 100,965/107,356 (94.0%) | 1 (ref) | | 1 (ref) | |
| Omicron | 8,837/218,185 (4.1%) | 209,348/218,185 (95.9%) | 0.68 (0.66, 0.70) | <.001 | 0.65 (0.63, 0.67) | <.001 |
| Two doses | | | | | | |
| Delta | 1,849/50,897 (3.6%) | 49,048/50,897 (96.4%) | 1 (ref) | | 1 (ref) | |
| Omicron | 5,558/200,284 (2.8%) | 194,726/200,284 (97.2%) | 0.76 (0.73, 0.80) | <.001 | 0.98 (0.93, 1.04) | 0.58 |
| Three doses | | | | | | |
| Delta | 28/546 (5.1%) | 518/546 (94.9%) | 1 (ref) | | 1 (ref) | |
| Omicron | 1,630/69,584 (2.3%) | 67,954/69,584 (97.7%) | 0.46 (0.32, 0.66) | <.001 | 0.58 (0.40, 0.84) | 0.004 |
| **Restricted to Omicron** | | | | | | |
| None or only one dose | 8,837/218,185 (4.1%) | 209,348/218,185 (95.9%) | 1 (ref) | | 1 (ref) | |
| Two doses | 5,558/200,284 (2.8%) | 194,726/200,284 (97.2%) | 0.69 (0.66, 0.71) | <.001 | 0.77 (0.72, 0.82) | <.001 |
| Three doses | 1,630/69,584 (2.3%) | 67,954/69,584 (97.7%) | 0.58 (0.55, 0.61) | <.001 | 0.37 (0.35, 0.40) | <.001 |

Patients with Delta infection and 0–1 doses as referent:

| | **Not hospitalized** | | **Hospitalized** | | **Crude RR (95% CI)** | | **p value** | **Adjusted RR (95% CI)** | | **p value** |
|---|---|---|---|---|---|---|---|---|---|---|
| | Delta | Omicron | Delta | Omicron | Delta | Omicron | | Delta | Omicron | |
| None or one dose | 100,965/310,313 (32.5%) | 209,348/310,313 (67.5%) | 6,391/15,228 (42.0%) | 8,837/15,228 (58.0%) | 1 (ref) | 0.68 (0.66, 0.70) | <.001 | 1 (ref) | 0.65 (0.63, 0.67) | <.001 |
| Two doses | 49,048/243,774 (20.1%) | 194,726/243,774 (79.9%) | 1,849/7,407 (25.0%) | 5,558/7,407 (75.0%) | 0.61 (0.58, 0.64) | 0.47 (0.45, 0.48) | . | 0.51 (0.47, 0.55) | 0.50 (0.47, 0.54) | . |
| Three doses | 518/68,472 (0.8%) | 67,954/68,472 (99.2%) | 28/1,658 (1.7%) | 1,630/1,658 (98.3%) | 0.86 (0.60, 1.24) | 0.39 (0.37, 0.42) | . | 0.42 (0.29, 0.62) | 0.24 (0.23, 0.26) | . |

**eTable 3. Relative risks of death by diagnosis with COVID-19 during periods of Omicron compared with Delta predominance, overall and according to vaccination status, New York City, August 2021–January 2022.**

| | Death | No death | Crude RR (95% CI) | p value | Adjusted RR (95% CI) | p value |
|---|---|---|---|---|---|---|
| **Variant** | | | | | | |
| Delta | 1,196/158,799 (0.8%) | 157,603/158,799 (99.2%) | 1 (ref) | | 1 (ref) | |
| Omicron | 2,696/488,053 (0.6%) | 485,357/488,053 (99.4%) | 0.66 (0.49, 0.89) | .006 | 0.81 (0.58, 1.13) | .22 |
| **By vaccination status** | | | | | | |
| None or only one dose | | | | .11 | | <.001 |
| Delta | 890/107,356 (0.8%) | 106,466/107,356 (99.2%) | 1 (ref) | | 1 (ref) | |
| Omicron | 1,534/218,185 (0.7%) | 216,651/218,185 (99.3%) | 0.85 (0.78, 0.92) | <.001 | 0.68 (0.63, 0.74) | <.001 |
| Two doses | | | | | | |
| Delta | 301/50,897 (0.6%) | 50,596/50,897 (99.4%) | 1 (ref) | | 1 (ref) | |
| Omicron | 865/200,284 (0.4%) | 199,419/200,284 (99.6%) | 0.73 (0.64, 0.83) | <.001 | 1.09 (0.95, 1.25) | .22 |
| Three doses | | | | | | |
| Delta | 5/546 (0.9%) | 541/546 (99.1%) | 1 (ref) | | 1 (ref) | |
| Omicron | 297/69,584 (0.4%) | 69,287/69,584 (99.6%) | 0.47 (0.19, 1.12) | .09 | 0.72 (0.27, 1.93) | .52 |
| **Restricted to Omicron** | | | | | | |
| None or only one dose | 1,534/218,185 (0.7%) | 216,651/218,185 (99.3%) | 1 (ref) | | 1 (ref) | |
| Two doses | 865/200,284 (0.4%) | 199,419/200,284 (99.6%) | 0.61 (0.57, 0.67) | <.001 | 0.82 (0.69, 0.98) | .03 |
| Three doses | 297/69,584 (0.4%) | 69,287/69,584 (99.6%) | 0.61 (0.54, 0.69) | <.001 | 0.39 (0.31, 0.47) | <.001 |

Patients with Delta infection and 0–1 doses as referent:

| | **No death** | | **Death** | | **Crude RR (95% CI)** | | **p value** | **Adjusted RR (95% CI)** | | **p value** |
|---|---|---|---|---|---|---|---|---|---|---|
| | Delta | Omicron | Delta | Omicron | Delta | Omicron | | Delta | Omicron | |
| None or one dose | 106,466/323,117 (32.9%) | 216,651/323,117 (67.1%) | 890/2,424 (36.7%) | 1,534/2,424 (63.3%) | 1 (ref) | 0.85 (0.78, 0.92) | .11 | 1 (ref) | 0.68 (0.63, 0.74) | <.001 |
| Two doses | 50,596/250,015 (20.2%) | 199,419/250,015 (79.8%) | 301/1,166 (25.8%) | 865/1,166 (74.2%) | 0.71 (0.63, 0.81) | 0.52 (0.47, 0.57) | . | 0.51 (0.42, 0.63) | 0.56 (0.46, 0.68) | . |
| Three doses | 541/69,828 (0.8%) | 69,287/69,828 (99.2%) | 5/302 (1.7%) | 297/302 (98.3%) | 1.10 (0.46, 2.65) | 0.51 (0.45, 0.59) | . | 0.36 (0.13, 0.99) | 0.26 (0.21, 0.33) | . |

**eFigure 1.** Eligibility for secondary analysis of Delta and Omicron infections based on whole-genome sequencing result.

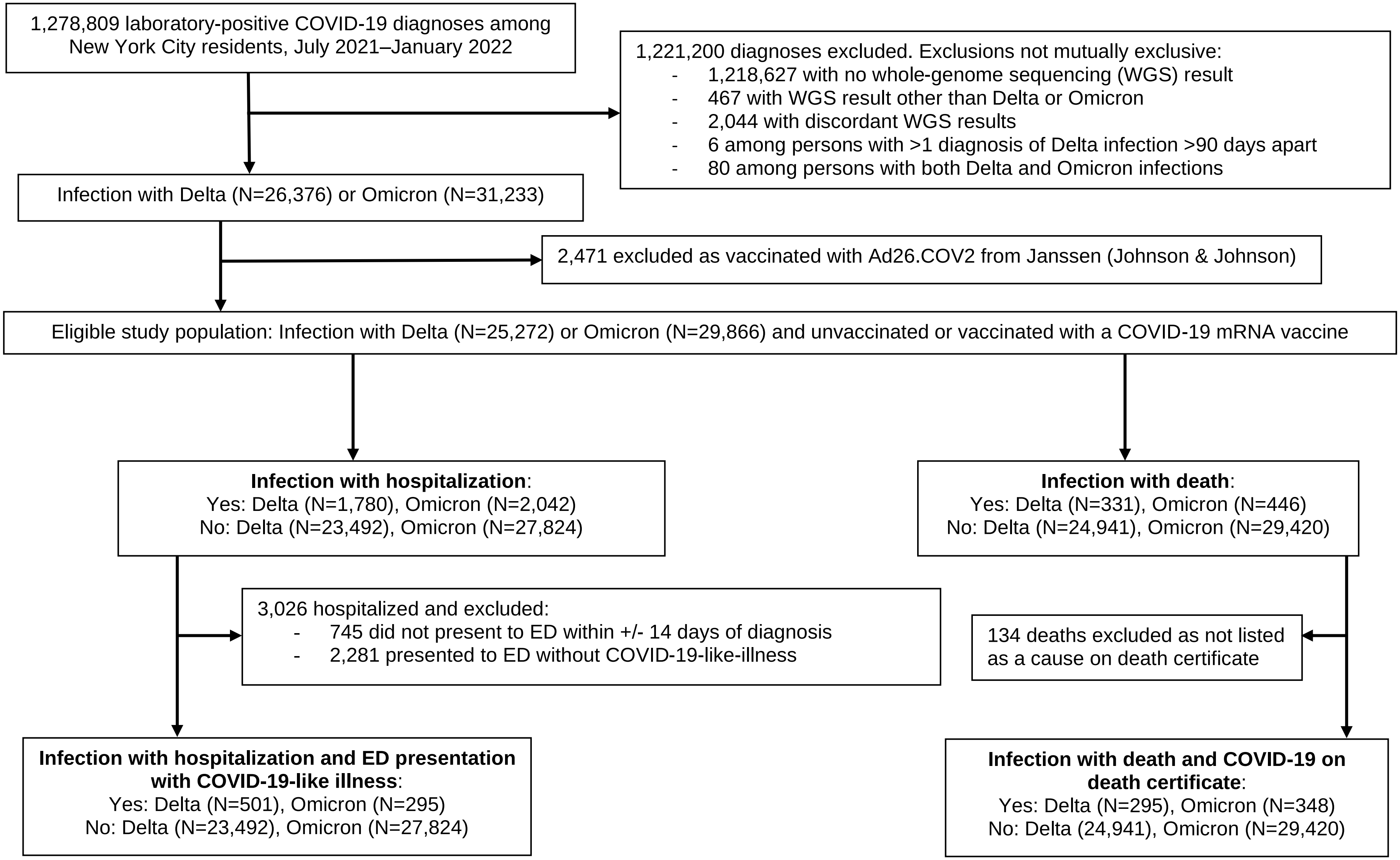

**eFigure 2. Relative risks of (A) hospitalization with COVID-19-like illness presentation and (B) death with COVID-19 indicated on the death certificate, by diagnosis during periods of Omicron compared with Delta predominance overall and according to vaccination status, New York City, August 2021–January 2022.**

(A) Hospitalization with COVID-19-like illness presentation

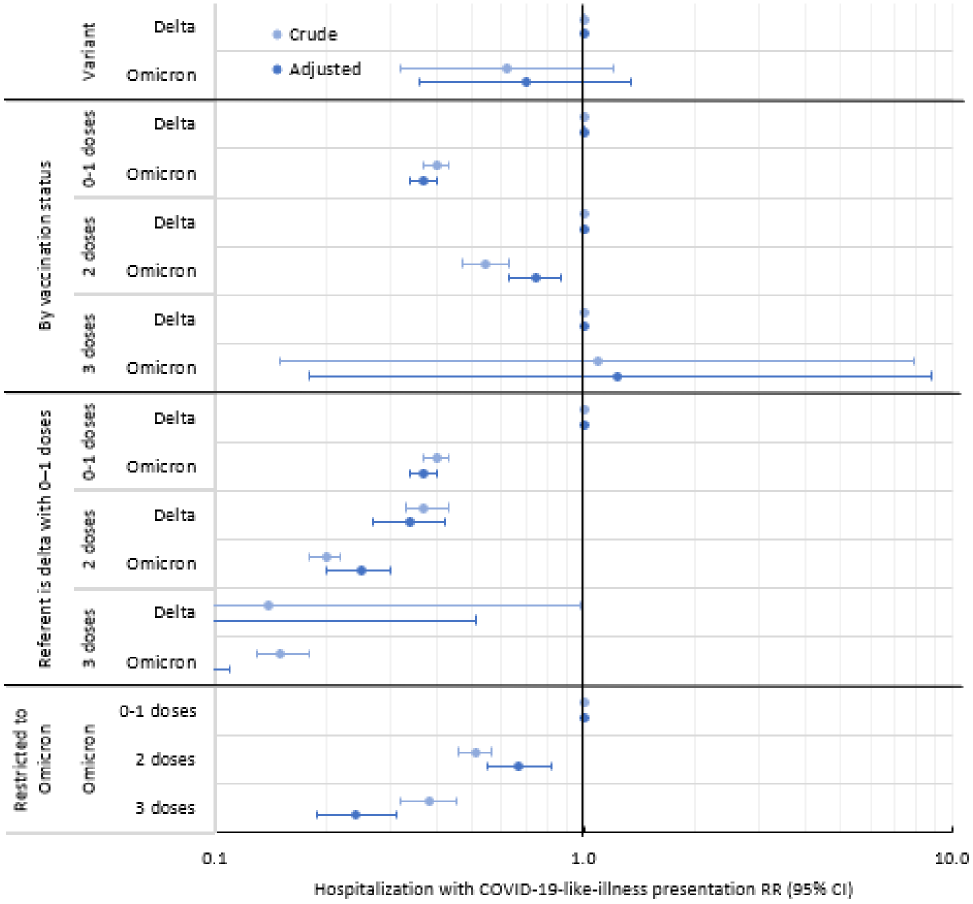

(B) Death with COVID-19 indicated on the death certificate

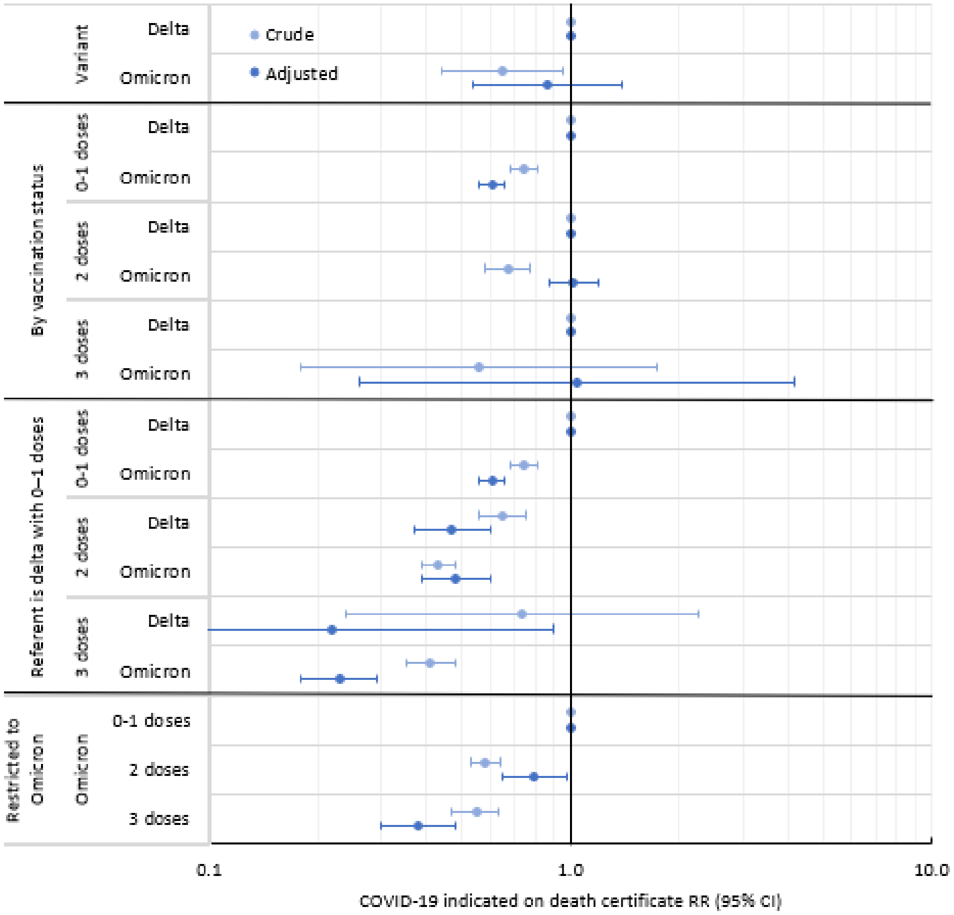

**eFigure 3. Relative risks of COVID-19 hospitalization and death for patients with Omicron compared with Delta sequencing results, overall and according to vaccination status, New York City, July 2021–January 2022.**

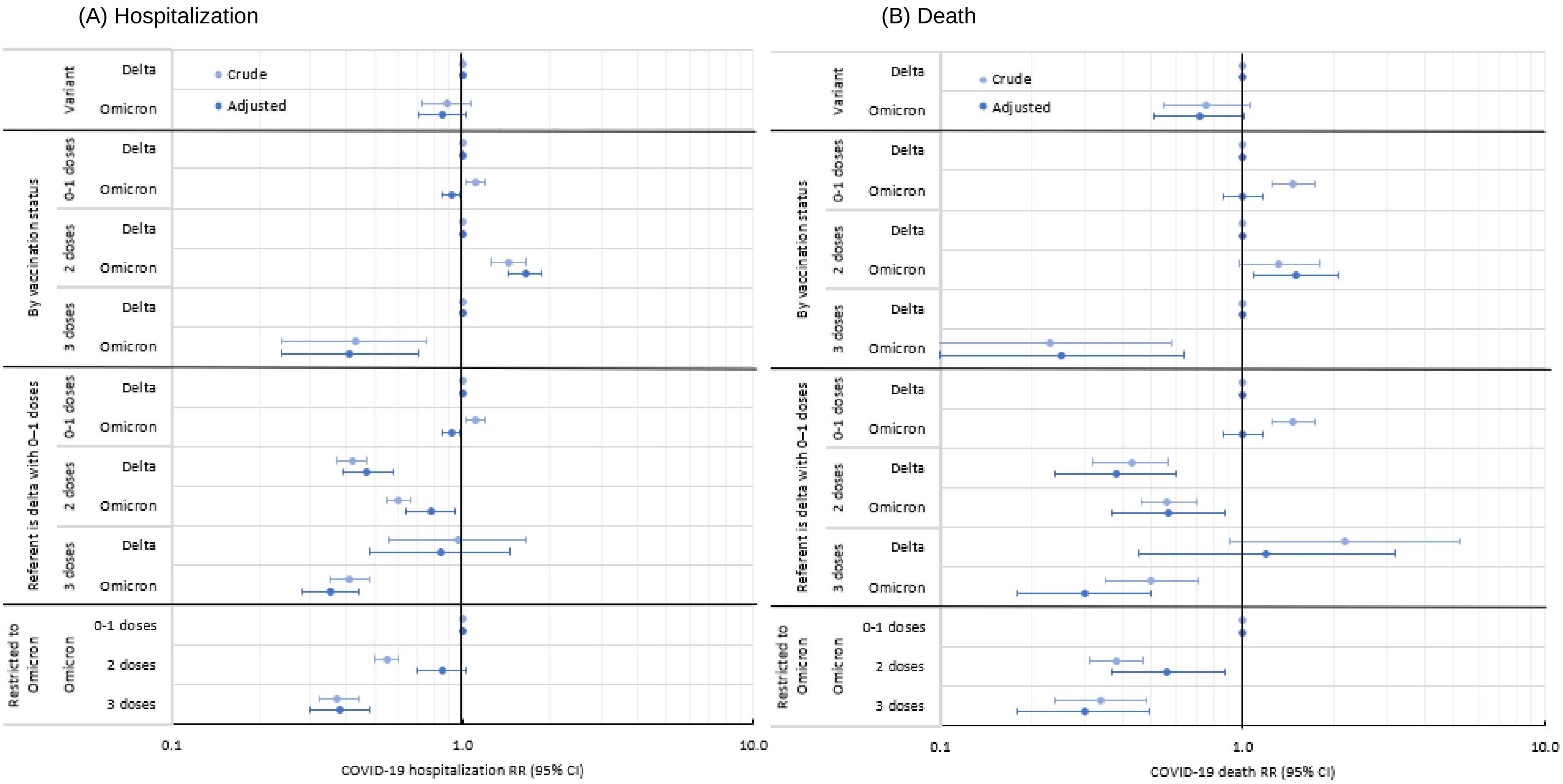
